# Impact of changing case definitions for COVID-19 on the epidemic curve and transmission parameters in mainland China

**DOI:** 10.1101/2020.03.23.20041319

**Authors:** Tim K. Tsang, Peng Wu, Yun Lin, Eric H. Y. Lau, Gabriel M. Leung, Benjamin J. Cowling

**Author notes:** **Corresponding author:** Peng Wu, School of Public Health, Li Ka Shing Faculty of Medicine, The University of Hong Kong, 7 Sassoon Road, Pokfulam, Hong Kong.

## Abstract

**Background:** When a new infectious disease emerges, appropriate case definitions are important for clinical diagnosis and also for public health surveillance. Tracking case numbers over time allows us to determine speed of spread and the effectiveness of interventions. Changing case definitions during an epidemic can affect these inferences.

**Methods:** We examined changes in the case definition for COVID-19 in mainland China during the first epidemic wave. We used simple models assuming exponential growth and then exponential decay to estimate how changes in the case definitions affected the numbers of cases reported each day. We then inferred how the epidemic curve would have appeared if the same case definition had been used throughout the epidemic.

**Findings:** From January through to early March 2020, seven versions of the case definition for COVID-19 were issued by the National Health Commission in China. As of February 20, there were 55,508 confirmed cases reported in mainland China. We estimated that when the case definitions were changed from version 1 to 2, version 2 to 4 and version 4 to 5, the proportion of infections being detected as cases were increased by 7.1-fold (95% credible interval (CI): 4.8, 10.9), 2.8-fold (95% CI: 1.9, 4.2) and 4.2-fold (95% CI: 2.6, 7.3) respectively. If the fifth version of the case definition had been applied throughout the outbreak, we estimated that by February 20 there would have been 232,000 (95% CI: 161,000, 359,000) confirmed cases.

**Interpretation:** The case definition was initially narrow, but was gradually broadened to allow detection of more cases as knowledge increased, particularly milder cases and those without epidemiological links to Wuhan or other known cases. This should be taken into account when making inferences on epidemic growth rates and doubling times, and therefore on the reproductive number, to avoid bias.

**Funding:** Commissioned grant from the Health and Medical Research Fund, Food and Health Bureau, Government of the Hong Kong Special Administrative Region.

## INTRODUCTION

When a newly emerging infectious disease is first identified, specifying appropriate case definitions can help to identify infected persons in an efficient manner [1]. Often a hierarchy of case definitions will be used, so that a “suspected case” could be defined based on some broad epidemiological and clinical criteria, for example patients with particular exposures or in particular geographic locations, with particular signs or symptoms, at a particular time. A “confirmed case” could be defined as a suspected case in which the pathogen of interest is identified or isolated using a specific laboratory test. Epidemiological and clinical information on patients who meet a case definition can inform the source(s) of infections, potential modes of transmission, transmission dynamics, impact and severity of the infection. All of this information is important for determining the optimal control measures.

COVID-19 is caused by the novel severe acute respiratory syndrome coronavirus 2 (SARS-CoV-2). It was first identified in a cluster of patients with atypical pneumonia in Wuhan in December 2019 [2]. Some of the patients had epidemiological links to the Huanan Seafood Wholesale Market in central Wuhan, and evidence provided by local health authorities suggested that most infections were from a zoonotic source with a limited amount of onwards human-to-human transmission [3]. During the latter part of January 2020 it became clear that infection was spreading efficiently from person to person, and then also that there was a broader clinical spectrum of infections [4]. As a consequence of the evolving information on epidemiological and clinical spectrum of infections, there have been a number of revisions to the case definition for COVID-19 in mainland China.

Here, we reviewed the various COVID-19 case definitions that have been used in mainland China as of 13 March 2020, and we examined the implications of changes in case definitions on our understanding of the epidemiology of COVID-19.

## METHODS

### Sources of data

We obtained the guidelines on diagnosis and treatment of COVID-19. The first two editions have not been released publicly while the third edition onwards have been released by the National Health Commission [5]. All of the original guidelines were published in Chinese as official government documents, and were translated into English by two authors (PW and YL). Epidemic curves by onset date and report date from December 2, 2019 to February 20, 2020 in China were extracted from the figures presented in the report of the WHO-China Joint Mission in February 2020 [6].

### Statistical analysis

We reviewed the various case definitions and highlighted the key changes in sequential updates. We fitted an exponential growth model to the incidence of cases to quantify the effect of changing case definitions on the epidemic curve. In the model, we assumed that each change in case definition increased the proportion of cases that would be detected among all infections. To account for the control measures, such as the lockdown in Wuhan and other cities in China on January 23, 2020 and the subsequent days [7], we allowed the growth rate to change on January 23. Because the interventions acted to prevent infections but the epidemic curve was based on date of symptom onset in our analysis, the effect of the interventions would be expected to have a slightly delayed effect on the epidemic curve which we accounted for by incorporating the incubation period distribution. The incubation period was assumed to follow a log-normal distribution with mean 5.2 days and standard deviation 3.9 days [8].

When changing the case definition, there could be a backfill of cases that fulfilled the new case definition around the change time. We allowed for backfill up to 10 days before each change in case definitions by assuming that a change in case definitions may have a partial effect on incidence before the change date *t*. For each day *k* before the change date *t*, this partial effect was assumed to be the probability that the cases at day *k* would be reported at or after the change date *t*. We assumed this onset-to-reporting distribution followed a Gamma distribution and inferred this distribution from the time series of onset and reporting in China by using the convolution of the symptom onset time series and the onset-to-reporting distribution to compute the expected number of reported cases for each day in the epidemic. Then we used a normal likelihood to the logarithm of observed and expected time series of cases by reporting date. We estimated the growth rate as one of the model parameters, and we estimated the doubling time by log(2) divided by the estimated growth rate. We fitted separate models for (1) Wuhan, (2) Hubei province excluding Wuhan, and (3) the rest of mainland China excluding Hubei province, to account for the regional differences in epidemic timing and potential transmissibility. We estimated the basic reproductive number *R*_*0*_, corresponding to the average number of secondary infections from one case at the start of the outbreak, using the formula 1/M(-r) [9], where r was the growth rate and M(.) was the moment generating function of the generation time distribution. We assumed the generation time distribution followed the same gamma distribution as a previously estimated serial interval distribution with mean 7.5 days and standard deviation 3.4 days [8]. We conducted a sensitivity analysis allowing backfill for up to 15 days prior to each change in case definitions.

To account for the uncertainty in estimates of the onset-to-reporting interval, and to allow us to quantify the uncertainty in model parameters including the growth rates, we performed our analysis in a Bayesian framework and constructed a Markov Chain Monte Carlo algorithm [10] that allowed joint parameter estimation. All statistical analyses were conducted using R version 3.5.2 (R Foundation for Statistical Computing, Vienna, Austria).

## RESULTS

Prior to January 15 we were unable to identify the case definition that was used in Wuhan to identify the earliest 41 confirmed cases. The first national guideline for diagnosis and treatment was issued on 15 January, and required six specific criteria to be met for a patient to be a confirmed case of COVID-19 (Table 1, Figure 1). Notably, patients needed to have an epidemiological link to Wuhan or a wet market in Wuhan and had to fulfil four clinical conditions indicative of viral pneumonia to be identified as suspected cases, and then had to have a respiratory specimen tested by full genome sequencing that showed a close homology with SARS-CoV-2 for the final confirmation of COVID-19. In the following days and weeks, a number of revisions were made to the case definitions, allowing gradually greater sensitivity in the criteria required for case confirmation (Figure 1).

**Table 1.** Definitions of suspected and confirmed cases of COVID-19 in seven versions of the National Guideline for Diagnosis and Treatment of the Novel Coronavirus Infection implemented in China since 15 January 2020.

| Version of guideline | Case definitions | Difference from the previous version | Date of issue | Date of online publication* |
| --- | --- | --- | --- | --- |
| 1 | **Medically observed case:**<br>Meet one of the epidemiological criteria and all the clinical manifestations below:<br>1. Epidemiological history:<br>(1) Travelled to Wuhan within two weeks before illness onset<br>(2) Had direct/indirect exposure to markets (especially wet markets) in Wuhan within two weeks before illness onset.<br>2. Clinical manifestations:<br>(1) Fever;<br>(2) Pneumonia indicated by chest radiograph;<br>(3) Reduced or normal white blood cell count, or reduced lymphocyte count during the early stage of the illness;<br>(4) After initiation of a standard antimicrobial treatment (following the "Guidelines for the Diagnosis and Treatment of Community-acquired Pneumonia in Chinese Adults (2016 Edition)" issued by Chinese Thoracic Society and the "Guidelines for the Diagnosis and Treatment of Community-acquired Pneumonia in Children (2019 Edition)" issued by National Health Commission) for 3 days, the patient's clinical conditions had not been significantly improved or become deteriorated.<br>**Confirmed case:**<br>A medically observed case has viruses isolated from respiratory specimens (sputum, throat swab, etc.) showing high toto a known novel coronavirus by the whole genome sequencing. | NA | Jan 15 | Not published online |
| 2 | **Suspected case** (previously named medically observed case):<br>Meet one of the epidemiological criteria and all the clinical manifestations below:<br>1. Epidemiological history<br>(1) travelled to or had lived in Wuhan within two weeks before illness onset;<br>(2) had a contact within 14 days before onset with a person showing fever and respiratory symptoms from Wuhan; | 1. Medically observed cases were renamed as suspected cases;<br>2. Removed an exposure to markets in Wuhan from the epidemiological criteria.<br>3. Added contact with symptomatic | Jan 18 | Not published online |
|  | (3) had a clustering occurrence.<br>2. Clinical manifestations:<br>(1) Fever;<br>(2) Pneumonia indicated by chest radiograph;<br>(3) Reduced or normal white blood cell count, or reduced lymphocyte count during early onset;<br>**Confirmed case:**<br>A suspected case has a detection of nucleic acid of the novel coronavirus from respiratory specimens including sputum, throat swab or lower respiratory tract specimens using the real-time fluorescent RT-PCR to test; or has isolated viruses showing high homology with a known SARS-CoV-2 by the whole genome sequencing. | persons from Wuhan as one of the epidemiological criteria;<br>4. Added clustering occurrence as one of the epidemiological criteria;<br>5. Removed failure in antibiotic treatment from the clinical manifestations;<br>6. Added RT-PCR as a laboratory test to confirm the infection of SARS-CoV-2. |  |  |
| 3 | **Suspected case** (previously named medically observed case):<br>Meet one of the epidemiological criteria and all the clinical manifestations below:<br>1. Epidemiological history:<br>(1) travelled to or had lived in Wuhan within 2 weeks before illness onset;<br>(2) had contacted patients from Wuhan who had fever and respiratory symptoms within 14 days before onset;<br>(3) had a clustering occurrence.<br>2. Clinical manifestations:<br>(1) Fever;<br>(2) Pneumonia indicated by chest radiograph;<br>(3) Low or normal white blood cell count, or low lymphocyte count during early onset;<br>**Confirmed case:**<br>A suspected case has a detection of nucleic acid of the novel coronavirus from respiratory specimens including sputum, throat swab or lower respiratory tract specimens using the real-time fluorescent RT-PCR to test; or has isolated viruses showing high homology with a known SARS-CoV-2 by the whole genome sequencing. | No difference in definitions for suspected and confirmed cases | Jan 22 | Jan 23 |
| 4 | **Suspected case:**<br>Meet one of the epidemiological criteria AND two of the clinical manifestations:<br>1. Epidemiological history:<br>(1) travelled to or had lived in Wuhan or other areas with sustained local transmission of COVID-19 within 14 days before illness onset; | 1. Expanded the epidemiological criteria by adding travel history to or residence in other areas than Wuhan having sustained local transmission of COVID-19 within 14 | Jan 27 | Jan 27 on the NHC website and Jan 28 on Chinese CDC website |
|  | (2) Contacted with patient(s) with fever or respiratory symptoms from Wuhan or other areas with sustained local transmission of COVID-19 within 14 days before illness onset;<br>(3) Had clustering occurrence, or epidemiologically linked to people infected with the SARS-CoV-2.<br>2. Clinical manifestations:<br>(1) Fever;<br>(2) Pneumonia indicated by chest radiograph;<br>(3) Low or normal white blood cell count, or low lymphocyte count during early onset;<br>**Confirmed case:**<br>A suspected case with one of the following etiological confirmations:<br>1. Respiratory or blood specimens tested positive for the nucleic acid of SARS-CoV-2 by the real-time fluorescent RT-PCR;<br>2. Viruses isolated from respiratory or blood specimens showing high homology with a known SARS-CoV-2 by whole genome sequencing. | days before onset.<br>2. Expanded the epidemiological criteria by adding contact history with persons having fever or respiratory symptoms from other areas than Wuhan having sustained local transmission of COVID-19 within 14 days before onset.<br>3. Revised diagnosis of suspected cases with 2 out of 3 clinical manifestations and one epidemiological criterion.<br>4. Specified the types of specimens used for laboratory tests. |  |  |
| 5 | **<u>Provinces outside Hubei:</u>**<br>**Suspected case:**<br>Meet one of the epidemiological criteria AND two of the clinical manifestations, OR meet all three clinical manifestations if without epidemiological history:<br>1. Epidemiological history:<br>(1) Travelled to or had lived in Wuhan or the surrounding areas or other communities with reported COVID-19 cases within 14 days before illness onset;<br>(2) Contacted with patient(s) infected with SARS-CoV-2 (positive for SARS-CoV-2 nucleic acid) within 14 days before onset;<br>(3) Contacted patients with fever or respiratory symptoms from Wuhan or the surrounding areas, or from communities with reported COVID-19 cases, within 14 days before illness onset;<br>(4) Had clustering occurrence.<br>2. Clinical manifestations:<br>(1) Fever and/or respiratory symptoms;<br>(2) Pneumonia indicated by chest radiograph;<br>(3) Low or normal white blood cell count, or low lymphocyte count during early onset;<br>**Confirmed case:**<br>A suspected case with one of the following etiological confirmations: | 1. Separate the case definitions for Hubei province from the rest of the country.<br>2. Added a new diagnostic type of cases, clinically confirmed case, specifically for Hubei province.<br>3. Added visiting/living in the surrounding areas of Wuhan or other areas having COVID-19 cases reported within 2 weeks before symptom onset as one of the epidemiological criteria;<br>4. Specified “having epidemiological link with a COVID-19 patient” as “contact a confirmed COVID-19 patient (positive for SARS-CoV-2 nucleic acid) within 14 days before onset”.<br>5. Added a suspected case being diagnosed without having an epidemiological criterion but | Feb 4 | Feb 5 |
|  | 1. Respiratory or blood specimens tested positive for the nucleic acid of SARS-CoV-2 by the real-time fluorescent RT-PCR;<br>2. Viruses isolated from respiratory or blood specimens showing high homology with a known SARS-CoV-2 by whole genome sequencing.<br>**<u>Hubei Province:</u>**<br>**Suspected case:**<br>Meet the clinical manifestations with or without an epidemiological criterion.<br>1. Epidemiological history:<br>(1) Travelled to or had lived in Wuhan or the surrounding areas or other communities with reported COVID-19 cases within 14 days before illness onset;<br>(2) Contacted with patient(s) infected with SARS-CoV-2 (positive for SARS-CoV-2 nucleic acid) within 14 days before onset;<br>(3) Contacted patients with fever or respiratory symptoms from Wuhan or the surrounding areas, or from communities with reported COVID-19 cases, within 14 days before illness onset;<br>(4) Had clustering occurrence.<br>2. Clinical manifestations:<br>(1) Fever and/or respiratory symptoms;<br>(2) Low or normal white blood cell count, or low lymphocyte count during early onset.<br>**Clinically diagnosed case:**<br>A suspected case with pneumonia indicated by chest radiograph.<br>**Confirmed case:**<br>A clinically diagnosed case or suspected case with one of the following etiological confirmations:<br>1. Respiratory or blood specimens tested positive for the nucleic acid of SARS-CoV-2 by the real-time fluorescent RT-PCR;<br>2. Viruses isolated from respiratory or blood specimens showing high homology with a known SARS-CoV-2 by whole genome sequencing. | fulfilling all three clinical criteria for patients outside of Hubei province.<br>6. Suspected cases in Hubei province could be diagnosed with only 2 of the clinical manifestations if without any epidemiological link while in other provinces a suspected cases had to fulfil 3 clinical criteria if without an epidemiological link. |  |  |
| 6 | **Suspected case:**<br>Meet one of the epidemiological criteria AND two of the clinical manifestations, OR meet all three clinical manifestations if without epidemiological history:<br>1. Epidemiological history:<br>(1) Travelled to or had lived in Wuhan or the surrounding areas or other communities | 1. Unified the case definitions for Hubei and other provinces by removing the clinically diagnosed case;<br>2. Provided suggestions that sputum | Feb 18 | Feb 19 |
|  | with reported COVID-19 cases within 14 days before illness onset;<br>(2) Contacted with patient(s) infected with SARS-CoV-2 (positive for SARS-CoV-2 nucleic acid) within 14 days before onset;<br>(3) Contacted patients with fever or respiratory symptoms from Wuhan or the surrounding areas, or from communities with reported COVID-19 cases, within 14 days before illness onset;<br>(4) Had clustering occurrence.<br>2. Clinical manifestations:<br>(1) Fever and/or respiratory symptoms;<br>(2) Pneumonia indicated by chest radiograph;<br>(3) Low or normal white blood cell count, or low lymphocyte count during early onset;<br>**Confirmed case:**<br>A suspected case with one of the following etiological confirmations:<br>1. Respiratory or blood specimens tested positive for the nucleic acid of SARS-CoV-2 by the real-time fluorescent RT-PCR;<br>2. Viruses isolated from respiratory or blood specimens showing high homology with a known SARS-CoV-2 by whole genome sequencing. | and lower respiratory specimens were preferred for PCR testing in the section of Laboratory Tests whereas removed the description of types of specimen to be collected for etiological tests in case definitions for confirmed cases. |  |  |
| 7 | **Suspected case:**<br>Meet one of the epidemiological criteria AND two of the clinical manifestations, OR meet all three clinical manifestations if without epidemiological history:<br>1. Epidemiological history:<br>(1) Travelled to or had lived in Wuhan or the surrounding areas or other communities with reported COVID-19 cases within 14 days before illness onset;<br>(2) Contacted with patient(s) infected with SARS-CoV-2 (positive for SARS-CoV-2 nucleic acid) within 14 days before onset;<br>(3) Contacted patients with fever or respiratory symptoms from Wuhan or the surrounding areas, or from communities with reported COVID-19 cases, within 14 days before illness onset;<br>(4) Had clustering occurrence ($\geq 2$ cases with fever and/or respiratory symptoms in a small area such as a family, an office, a school class, etc., within 2 weeks).<br>2. Clinical manifestations:<br>(1) Fever and/or respiratory symptoms;<br>(2) Pneumonia indicated by chest radiograph;<br>(3) Low or normal white blood cell count, or low lymphocyte count during early onset; | 1. Added details to define a clustering occurrence;<br>2. Added serologic tests as one of the etiological confirmation approaches. | Mar 3 | Mar 4 on NHC website |
|  | **Confirmed case:**<br>A suspected case with one of the following etiological confirmations:<br>1. Respiratory or blood specimens tested positive for the nucleic acid of SARS-CoV-2 by the real-time fluorescent RT-PCR;<br>2. Viruses isolated from respiratory or blood specimens showing high homology with a known SARS-CoV-2 by whole genome sequencing;<br>3. Serum tested positive for SARS-CoV-2 specific IgM and IgG antibodies; the serum SARS-CoV-2-specific IgG antibody test changed from negative (undetectable) to positive (detectable), or the IgG antibody level is 4 times higher in the convalescent serum than in the serum collected at the acute phase of the infection. |  |  |  |

\*online publication refers to whether the document was available on the official websites of NHC and China CDC.

**Figure 1:**
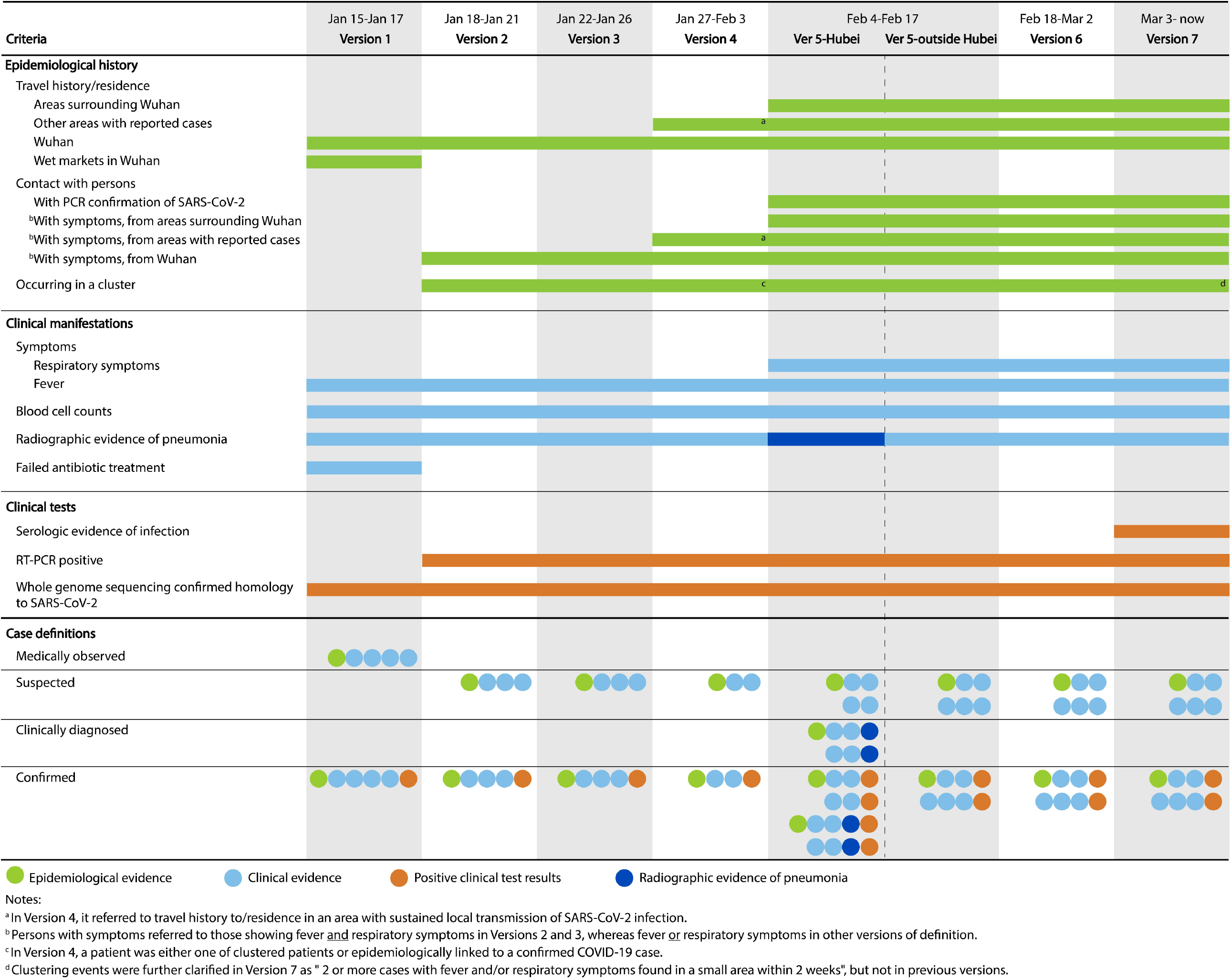
Evolution of the case definitions of COVID-19 in seven editions of the National Guideline for Diagnosis and Treatment of COVID-19 in mainland China since 15 January 2020.

The second edition of the case definitions removed the requirement for failure of antibiotic treatment, and allowed PCR confirmation in addition to whole genome sequencing. The third edition modified and clarified the definitions of “severe” and “critical” cases (not discussed further, here). The fourth edition allowed patients to have an epidemiological link to other areas with reported cases, instead of being restricted to Wuhan, and suspected cases required only two instead of all the three types of clinical manifestations in addition to an epidemiological link. Perhaps the greatest change was in the fifth edition, which introduced a new category of cases specifically for Hubei province which is the epicenter of the outbreak and had the largest number of cases identified in the country. Here, “clinically confirmed” cases were patients that met clinical criteria and had radiological evidence of pneumonia with or without certain epidemiological link but did not need to have a virological confirmation of infection. In the sixth edition, this criteria was removed and no distinction was made between cases inside or outside Hubei province. In the seventh edition, serology was added as an additional option for laboratory confirmation.

We modelled the effects of changes in case definition from version 1 to version 2, from version 2 to 4, and from version 4 to 5. We did not explore the effects of changing from version 2 to 3 since version 3 only included updates to the severity classifications. We were not able to explore the change after version 5 since we only analysed data up to 20 February which included just the first two days after the release of version 6. We were not able to find information on incidence of cases by illness onset date after February 20 and had to censor our analysis at that point.

The changes in case definitions had a clear impact on the proportion of infections that were identified and counted as confirmed cases. As of February 20, there were 55,508 confirmed cases in China, among which 26,927, 15,847 and 12,734 were from Wuhan, Hubei province excluding Wuhan, and China excluding Hubei province, respectively. We estimated that the mean onset-to-reporting delay was 8.6 days (95% CI: 7.4, 10.1) and the 95^th^ percentile of this distribution was 15.7 days (95% CI: 13.0, 20.1). Allowing for a 10 day backfill of cases, we estimated that when the case definitions were changed from version 1 to 2, version 2 to 4 and version 4 to 5, the proportion of infections being identified as COVID-19 cases were increased by 7.1-fold (95% credible interval (CI): 4.8, 10.9), 2.8-fold (95% CI: 1.9, 4.2) and 4.2-fold (95% CI: 2.6, 7.3) respectively (Figure 2).

**Figure 2:**
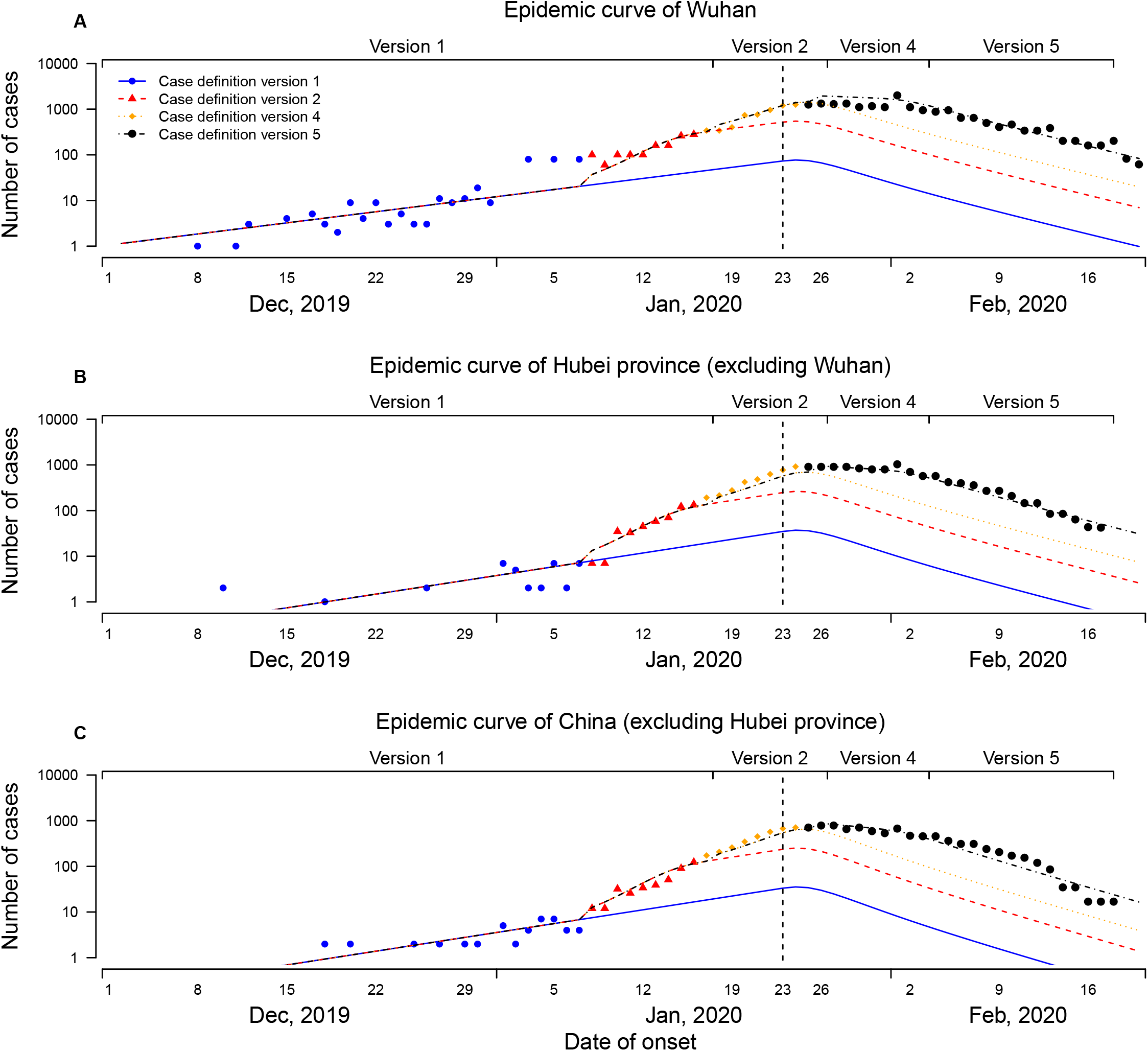
Number of reported COVID-19 cases by date of onset and the estimated daily numbers of cases in the hypothetical analyses by assuming a version of case definitions to be applied throughout the study period in mainland China, as of 20 February 2020. Symbols show daily numbers of cases reported, and colours indicated reported cases (symbols) or estimated cases (lines) at different time periods in line with the timeline when different versions of COVID-19 case definitions were released. The color for changing case definitions was adjusted earlier to reflect that assumption that there was a backfill of symptomatic cases who had not yet presented for diagnosis up to 10 days before each change in case definition, and therefore the effect of changing case definition would appear to modify the proportion of infections captured as cases before the actual day of change.

In a hypothetical analysis assuming that the case definitions from version 5 had been applied throughout the outbreak, and that laboratory testing with RT-PCR had been available from an early stage of the epidemic, we estimated that 232,000 (95% CI: 161,000, 359,000) cases would have met the case definition and could have been detected by February 20, of which 127,000 (95% CI: 86,000, 198,000), 55,000 (95% CI: 38,000, 86,000) and 50,000 (95% CI: 34,000, 78,000) cases were from Wuhan, Hubei excluding Wuhan, and China excluding Hubei respectively (Figure 3). In this same hypothetical analysis, among the 127,000 cases that we estimated in Wuhan by February 20, we estimated that there would have been 11,000 infections (95% CI: 7,000, 21,000) that met version 5 of the case definition with illness onset by January 1, 2020. In the observed data, there were 114 confirmed COVID-19 cases with illness onset by January 1, 2020, corresponding to around 1% of our estimated total. Prior January 23, we estimated that 92% (95% CrI: 88%, 95%) cases were undetected.

**Figure 3:**
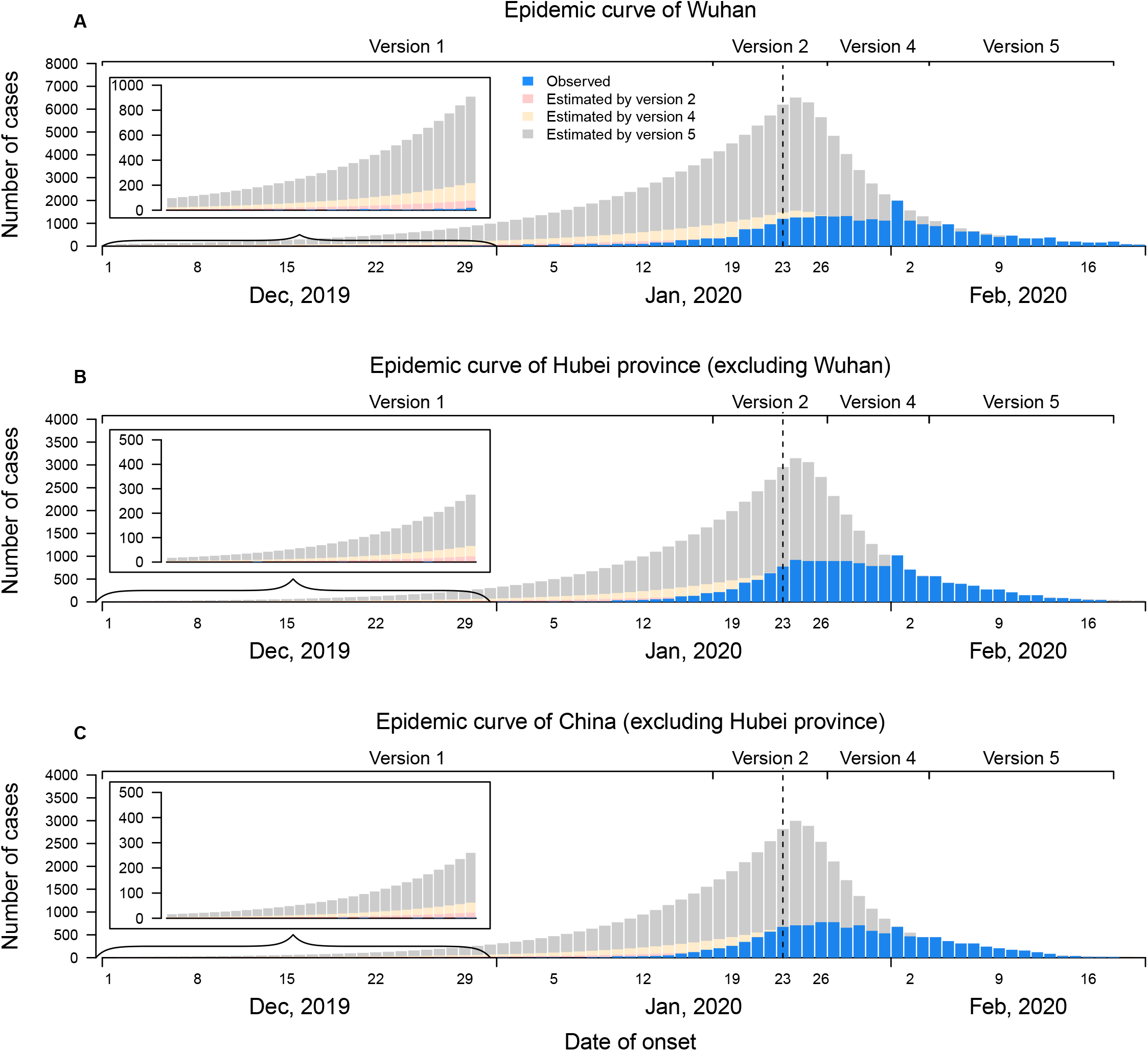
Observed (blue bars) and augmented occurrence by different case definition (red bars by case definition version 2, yellow bars by case definition version 4 and grey bars by case definition version 5) of COVID-19 cases by date of illness onset in Wuhan (Panel A), Hubei province excluding Wuhan (Panel B) and other provinces in mainland China excluding Hubei province (Panel C).

We estimated that after implementation of control measures on January 23 the growth rate declined substantially to below zero, from 0.08 to −0.15 in Wuhan which was a change of −0.23 (95% CI: −0.27, −0.20). The corresponding changes in growth rate were - 0.26 (95% CI: −0.30, −0.22) and −0.28 (95% CI: −0.32, −0.25) for Wuhan, Hubei excluding Wuhan, and for China excluding Hubei province, respectively. This suggested that the control measures were very effective, reducing the effective reproductive number to well below one.

After adjusting for the changes in case definitions, we estimated that the epidemic growth rate before January 23 was around 0.08 to 0.10 and the doubling time was around 7.0 to 8.7 days for these three geographic areas, and the differences among them were not statistically significant (Table 2). If instead the change in case definitions was unaccounted for, the growth rate would have been substantially overestimated and the doubling time would have been substantially underestimated (Table 2). Using a growth rate of 0.08-0.10 with a mean serial interval of 7.5 days [8] would lead to *R*_*0*_ estimates in the range 1.8-2.0. If we instead used the erroneous growth rate estimates of 0.15-0.19 (Table 2) we would obtain *R*_*0*_ estimates in the range of 2.8-3.5.

**Table 2.** Estimates of the epidemic growth rate and doubling time prior to 23 January 2020, with or without adjustment for changes in case definitions.

| **Parameter** | **Adjustment** | **Wuhan** | **Hubei province**<br>**excluding Wuhan** | **China excluding**<br>**Hubei province** |
| --- | --- | --- | --- | --- |
| Growth<br>rate, per<br>day | With adjustment for<br>changes in case<br>definitions | 0.08 (0.06, 0.10) | 0.10 (0.08, 0.12) | 0.10 (0.08, 0.12) |
|  | Without adjustment | 0.15 (0.14, 0.17) | 0.18 (0.13, 0.28) | 0.19 (0.16, 0.24) |
| Doubling<br>time, days | With adjustment for<br>changes in case<br>definitions | 8.7 (7.3, 10.8) | 7.0 (5.8, 8.8) | 7.0 (5.8, 8.7) |
|  | Without adjustment | 4.5 (4.1, 4.8) | 3.9 (2.4, 5.3) | 3.6 (2.9, 4.3) |

Notes:
<sup>a</sup> In Version 4, it referred to travel history to/residence in an area with sustained local transmission of SARS-CoV-2 infection.
<sup>b</sup> Persons with symptoms referred to those showing fever and respiratory symptoms in Versions 2 and 3, whereas fever or respiratory symptoms in other versions of definition.
<sup>c</sup> In Version 4, a patient was either one of clustered patients or epidemiologically linked to a confirmed COVID-19 case.
<sup>d</sup> Clustering events were further clarified in Version 7 as " 2 or more cases with fever and/or respiratory symptoms found in a small area within 2 weeks", but not in previous versions.

In a sensitivity analysis allowing for 15 days of backfill each time the case definition changed, the proportion of infections being identified as COVID-19 cases were increased by 3.0-fold to 8.8-fold. We estimated that 253,000 (95% CI: 158,000, 436,000) cases would have met the case definition and could have been detected by February 20. These estimates were slightly higher but as expected given the backfill period was longer.

## DISCUSSION

We found that changes in case definitions of COVID-19 in China led to stepwise increases in the proportion of all infections identified as cases, by 7.1-fold, 2.8-fold and 4.2-fold with updates to versions 2, 4 and 5 of the case definitions, respectively. Overall, we estimated that around 232,000 cases could have been confirmed in the first wave of COVID-19 in China by late February 2020 if, hypothetically, version 5 of the case definitions had been used throughout. Certainly, the number of infected persons is likely to be greater than 232,000 because many mild cases were not tested or confirmed, and some infections were asymptomatic [11]. We estimated that many cases were undetected when using earlier case definition, which is consistent with another recent study which estimated that around 85% cases were undetected prior to January 23, when case definition 2 was used [12].

The introduction of “clinically confirmed” cases in the fifth edition of the case definitions allowed a large number of highly suspected cases who could not receive a virologic test to be isolated and treated in time and reallocation of laboratory testing resources to be used for identifying and then isolating cases in the community as part of the containment efforts. This category was removed within a week, in the sixth edition of case definitions (Table 1) as explained by health officials that laboratory testing was sufficient to confirm all cases and the “clinically confirmed” category was unnecessary [13]. However confirmation of viral pneumonia with radiological evidence could be an important alternative for diagnosis and surveillance of COVID-19 in locations with limited laboratory testing capacity, and would also be a good option if or when a surge in COVID-19 consultations exceeds local laboratory capacity. This could be combined with testing a portion of the clinical confirmed cases to correct the actual case numbers afterward [14].

Case definitions are often developed for outbreak investigations in which the objective is to identify the source of infections [15] while only later if an epidemic occurs are case definitions used for surveillance. In the case of the COVID-19 epidemic in China, the initial case definitions for COVID-19 allowed investigation of potential animal exposures and infections epidemiologically linked with the epicentre, Wuhan, but might not capture cases linked with wider areas potentially affected by COVID-19 [16]. Similarly, the earlier case definitions had more specific requirements for clinical manifestations given the limited knowledge of the novel virus, leading to a low sensitivity for case identification including an under-detection of milder infections [16, 17]. As the evidence of the clinical spectrum of COVID-19 became available the case definition was rightly updated to account for this.

However, our analysis demonstrated that estimates of key epidemiological parameters using epidemic curves could be very biased if they do not account for such changes in case definitions. Specifically, we found that if we had estimated the exponential growth in the epidemic curve without accounting for the changes in case definitions, we would have substantially overestimated the growth rate and substantially underestimated the doubling time (Table 2). There are a number of high estimates of growth rates and *R*_*0*_ in the grey literature on preprint servers, some of which might suffer from this particular bias [18, 19]. Other high estimates of growth rates or *R*_*0*_ based on epidemic curves by reporting date might have overestimated transmissibility because of the shortening in onset-to-reporting delays as the epidemic progressed.

Our findings also suggest caution may be needed for analyses of the trajectories of epidemic curves elsewhere. Epidemics could appear to be growing faster than they actually are, because of rapid expansions in testing practices. The availability of and resolve for laboratory testing will also be a major factor shaping epidemic curves [20], which will be important to guide the public health responses. Due to the limited capacity for confirmation tests, Switzerland has announced it would stop testing mild cases and restrict such to those who are more ill [21], and other countries may adopt the same approach as case numbers increase. As discussed above, radiological confirmation could be a potential alternative to track the incidence of hospitalised cases.

A limitation of our study is that we did not formulate an individual-based mechanistic transmission model, but used a simple model with exponential growth and then exponential decay (Figure 2). Further work could explore more complex dynamic models, allowing for the marginal effects of different types of interventions that were introduced at different times towards the end of January 2020, in addition to accounting for the changes in case definitions. Analyses of the effects of interventions in China should be evaluated carefully if they do not account for the changes in case definitions. Second, we were only able to collect the data for the epidemic curve up to February 20, 2020 from published information. Therefore, we cannot evaluate the impact of changes in the case definition from version 5 to 6 and from version 6 to 7, although case numbers have been substantially declined after February 20.

In conclusion, we have shown that changes in case definitions had a very substantial effect on the proportion of all infections identified as cases as time progressed, and therefore also had a very substantial effect on the epidemic curve. Ignoring those changes would have led to biased estimates of some key epidemiological parameters. We estimated that there could have been 232,000 cases by February 20 if, hypothetically, version 5 of the case definitions had been used throughout the epidemic. However this would be an underestimate of the number of infections up to that point because it would not have captured some mild or asymptomatic cases. Serological studies could be used to reveal the true cumulative incidence of infections.

## Data Availability

We used publicly available information on confirmed case counts, and a spreadsheet can be obtained from the corresponding author on request.

## ACKNOWLEDGMENTS

The authors thank Julie Au for administrative support

